# Hyaluronan in COVID-19 morbidity, a bedside-to-bench approach to understand mechanisms and long-term consequences of hyaluronan

**DOI:** 10.1101/2023.02.10.23285332

**Authors:** Urban Hellman, Ebba Rosendal, Joakim Lehrstrand, Johan Henriksson, Tove Björsell, Max Hahn, Björn Österberg, Luiza Dorofte, Emma Nilsson, Mattias N.E. Forsell, Anna Smed-Sörensen, Anna Lange, Mats Karlsson, Clas Ahlm, Anders Blomberg, Sara Cajander, Ulf Ahlgren, Alicia Edin, Johan Normark, Anna K Överby, Annasara Lenman

## Abstract

**Background:** We have previously shown that lungs from deceased COVID-19 patients are filled with hyaluronan (HA). In this translational study, we investigated the role of HA in all stages of COVID-19 disease, to map the consequences of elevated HA in morbidity and identify the mechanism of SARS-CoV-2-induced HA production.

**Methods:** Lung morphology was visualized in 3D using light-sheet fluorescence microscopy. HA was verified by immunohistochemistry, and fragmentation was determined by gas-phase electrophoretic molecular mobility analysis. The association of systemic HA in blood plasma and disease severity was assessed in patients with mild (WHO Clinical Progression Scale, WHO-CPS, 1-5) and severe COVID-19 (WHO-CPS 6-9), during the acute and convalescent phases and related to lung function. *In vitro* 3D-lung models differentiated from primary human bronchial epithelial cells were used to study effects of SARS-CoV-2 infection on HA metabolism.

**Findings:** Lungs of deceased COVID-19 patients displayed reduced alveolar surface area compared to healthy controls. We verified HA in alveoli and showed high levels of fragmented HA both in lung tissue and aspirates. Systemic levels of HA were high during acute COVID-19 disease, remained elevated during convalescence and associated with reduced diffusion capacity. Transcriptomic analysis of SARS-CoV-2-infected lung models showed dysregulation of HA synthases and hyaluronidases, both contributing to increased HA in apical secretions. Corticosteroid treatment reduced inflammation and, also, downregulated HA synthases.

**Interpretation:** We show that HA plays a role in COVID-19 morbidity and that sustained elevated HA concentrations may contribute to long-term respiratory impairment. SARS-CoV-2 infection triggers a dysregulation of HA production, leading to increased concentrations of HA that are partially counteracted by corticosteroid treatment. Treatments directly targeting HA production and/or degradation can likely be used early during infection and may alleviate disease progression and prevent long-term lung complications.

## INTRODUCTION

The ongoing pandemic caused by severe acute respiratory syndrome coronavirus 2 (SARS-CoV-2) has hitherto caused more than 6.8 million reported deaths globally (by March 22, 2023, according to WHO). The clinical manifestations of COVID-19 range from asymptomatic or mild disease with symptoms from the upper respiratory tract to severe pneumonitis with acute respiratory distress syndrome (ARDS) and multiorgan failure. Several pathophysiological mechanisms have been described as contributing to respiratory failure in COVID-19, including hyperinflammation with disturbed coagulation, leading to disseminated pulmonary microthrombi as well as diffuse alveolar damage, alveolar septal fibrous proliferation and pulmonary consolidation (1, 2).

Hyaluronan (HA) is a glycosaminoglycan that constitutes an important structural component of the extracellular matrix in tissues. Through interactions with cell-surface receptors, HA also regulates cellular functions, such as cell-matrix signaling, cell proliferation, angiogenesis and cell migration (3). HA is often present in its high molecular weight structure in healthy tissue and has an anti-inflammatory effect (4). In contrast, low molecular weight HA has been shown to have a pro-inflammatory effect when HA fragments bind to toll-like receptors and induce NF-κB signaling (5). HA also influences the development of fibrosis by affecting fibroblast proliferation, differentiation, and motility (6).

In addition, HA has a very high water-binding capacity, with the ability to occupy large hydrated volumes up to 1,000 times its molecular mass, which can promote edema formation (3). Accumulation of HA is associated with ARDS (7), and recent publications have shown increased levels of HA in the lungs of deceased COVID-19 patients (8, 9) as well as elevated plasma levels of HA in severe cases (10, 11). Anti-inflammatory treatment with the corticosteroid dexamethasone results in lower mortality amongst hospitalized COVID-19 patients (12). Corticosteroids are known to be effective in reducing HA levels in other inflammatory syndromes (13, 14), thus, clearance of HA may be critical in disease resolution following COVID-19 (8). We recently found that severe COVID-19 is an important risk factor for impaired respiratory function, characterized by a decrease in diffusion capacity (DL_CO_), 3-6 months after the infection (15). However, if the lung function correlates with systemic HA is currently not clear.

Here we set out to investigate HA at all stages of COVID-19 disease, from acute infection to convalescence in mild, severely ill and fatal cases. Our aim was to understand molecular mechanisms involved in the pathological overproduction of HA in disease and pathogenicity. In addition, we established an *in vitro* 3D-lung model to investigate the impact of SARS-CoV-2 infection on HA metabolism as well as the role of corticosteroid treatment to define molecular disease mechanisms.

## RESULTS

### Morphological differences induced by COVID-19 visualized by light sheet fluorescent microscopy

We have previously shown that the lungs of fatal COVID-19 patients are filled with HA (8). However, it is not clear which consequences HA has on the structural integrity of the lung and the alveolar volume. To address this, we set out to morphologically determine the three-dimensional organization within lung biopsies with light sheet fluorescence microscopy (LSFM). These biopsies originated from three deceased COVID-19 patients, four healthy donors who underwent lung resection, and one patient who underwent lung resection ten weeks post intensive care treatment due to severe COVID-19 infection, here referred as recovered.

The lung biopsies were processed, cleared and subjected to LSFM visualizing the autofluorescence (figure 1A). Major differences in the morphology of the lung were detected by maximum intensity projection (figure 1B-D, supplementary figure 1, and supplementary movie 1-3). The lungs of fatal COVID-19 patients were considerably denser compared to the healthy controls and recovered lung. Optical sections of the tissue revealed thin alveolar walls in the controls and recovered, whereas the distances between the alveoli in COVID-19 lungs were larger (figure 1E-G). The size distribution of the alveoli diameter was also smaller in the COVID-19 compared to control and recovered lung (figure H). The number of empty alveoli was fewer in the COVID-19 lungs as visualized in the surface rendering of the alveolar space (figure 1I-K). Next, the surface of the alveoli was quantified to better understand the capacity of the gas exchange in the samples. The isosurfaces of the alveoli were determined and showed a major loss of alveoli surface in the COVID-19 lungs compared to the healthy controls (figure 1L). Worth noting is that the lung morphology found in the recovered COVID-19 patient showed high similarity to the healthy controls.

**Figure 1.**
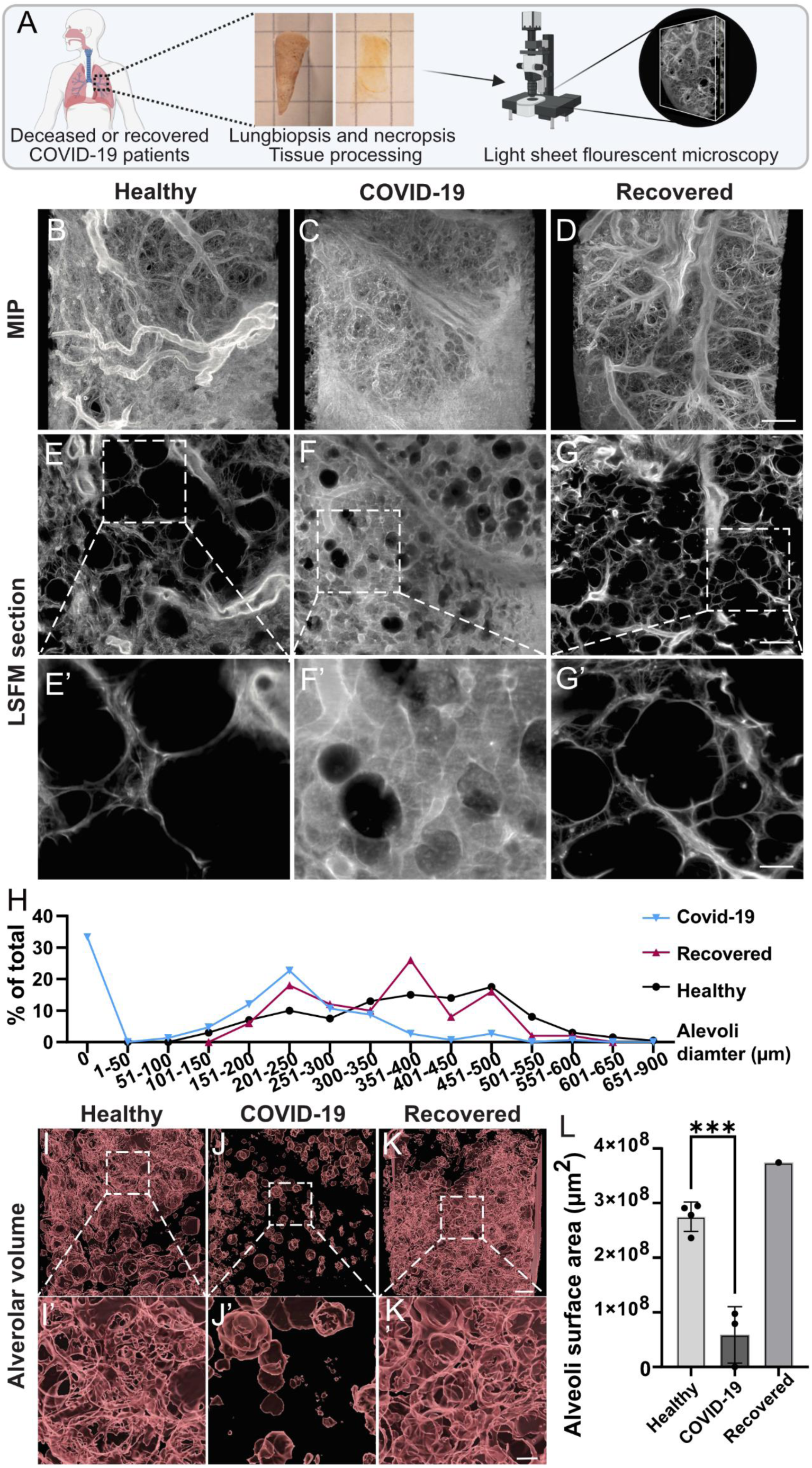
Light sheet fluorescent microscopy (LSFM) of biopsies from deceased COVID-19 patients displays a reduced alveolar surface. A) Schematic overview of LFSM procedure with tissue biopsies from healthy donors (n=4), deceased COVID-19 patients (n=3) and recovered COVID-19 patient (n=1). B-D) Maximum intensity projections of one representative biopsy from each patient group. Scale bar in D is 500 µm. E-G) LSFM sections and magnifications showing the alveolar structure in each patient group. Scale bar in G is 500 µm and scalebar in G’ is 100 µm. H) Size distribution of alveolar diameter measured in LSFM sections, ten random diameter measurements in five different z planes from each patient. I-K) Iso-surfaced empty space in the lung biopsies as a proxy for alveolar volume. Scalebar in K is 400 µm and scalebar in K’ is 100 µm. L) Quantification of the surface volume surrounding the empty space as a proxy for alveolar surface volume. Statistical significance was calculated by unpaired t-test (***p < 0.001).

To confirm the presence of HA in alveoli and to see if there is a difference in accumulation between the upper and lower lung lobes, histochemistry was performed on five COVID-19 necropsies (figure 2A-B). We did not observe a major difference between patients or lung regions, but extensive accumulation of HA was seen in COVID-19 necropsy compared to healthy controls (figure 2C). Collagen coils, a marker for irreversible lung damage, were also found in large areas of the necropsy where no remains of alveolar walls could be detected (figure 2C).

**Figure 2.**
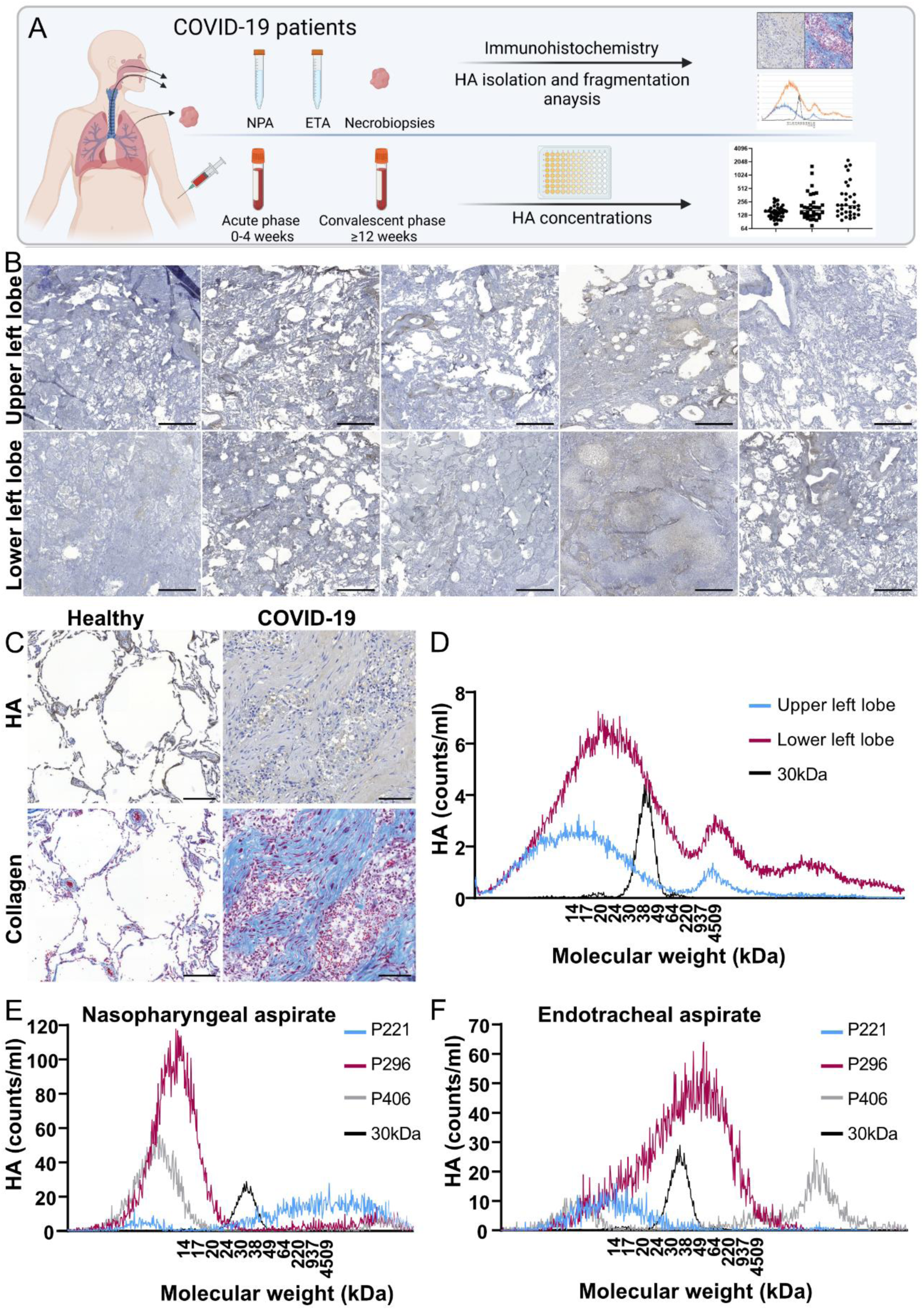
Lung necropsies and lung aspirates from severe COVID-19 patients show large amounts of hyaluronan (HA) and a high degree of HA fragmentation. A) Schematic overview of collection of patient samples for HA analysis. B) Lung biopsies from the upper and lower left lobe of five deceased COVID-19 patients were stained for HA. Scale bar is 1000 µm. C) Collagen and HA staining in lung biopsies from healthy lung tissue (scale bar is 200 µm) and from a deceased COVID-19 patient (scale bar is 100 µm). The COVID-19 lung biopsy shows areas with no alveoli structures. D) HA was isolated from the lung biopsies in B (n=5) and a size distribution analysis was performed with gas-phase electrophoretic mobility molecular analysis (GEMMA). Shown is an average of all five patients. High molecular weight HA with a known molecular size of 30 kDa was included as control. HA was also isolated and subjected to size analysis by GEMMA from E) nasopharyngeal aspirate and F) endotracheal aspirate collected from three severe COVID-19 patients and displayed individually for each patient.

### Severe COVID-19 induces fragmentation of HA in lung tissue and aspirates

As fragmented low molecular HA increases at sites of active inflammation and display a proinflammatory activity, HA fragmentation likely contributes to disease progression of COVID-19. HA was extracted from necropsies derived from the upper, and lower left lobe of five COVID-19 patients to investigate this. The isolated HA was subjected to size determination analysis by gas-phase electrophoretic mobility molecular analysis (GEMMA). Both the upper and lower lung lobes displayed highly fragmented HA (figure 2D). Although we did not observe an apparent difference in the abundance of HA with histochemistry between the upper and lower lung lobes (figure 2B), we found higher levels of both total HA and fragmented HA in the lower parts of the lung with GEMMA (figure 2D, area under the curve). Since fragmented HA induces a proinflammatory state and initiates migration of immune cells (16), we analysed the presence of neutrophils in lungs by staining for elastase. We detected major neutrophil infiltration in COVID-19 lungs compared to uninfected controls (supplementary figure 2). Next, nasopharyngeal aspirates (NPA) and endotracheal aspirates (ETA) collected from three COVID-19 patients were analysed to investigate the fragmentation process of HA during ongoing severe disease. All patients displayed fragmented HA, however, in contrast to the fragmentation in the necropsy samples, there were large individual variations between the total amount and degree of fragmentation (figure 2E-F). The finding that HA accumulated in the lungs and aspirates of severely diseased patients prompted us to measure the systemic levels of HA. We performed this in a cohort of mild and severely ill patients during acute disease and convalescence. Our objective was to investigate if plasma concentrations of HA could be used as a biomarker for disease severity.

### Hyaluronan levels in plasma increase in severe COVID-19 and correlate with a reduction in lung function at follow up

Patients with COVID-19 infection classified as severe, based on requirement of high-flow nasal oxygen treatment and/or admission to the intensive care unit (ICU) corresponding to WHO-CPS 6-10 (17), were matched according to age to patients with mild COVID-19 (WHO-CPS 1-5). In total 103 individuals were selected; 37 patients were classified as severe and 66 patients as mild. The demography and clinical characteristics are presented in table 1. The groups differed significantly in BMI, obesity, minimum levels of hemoglobin, maximum levels of C-reactive protein, white blood cell count, and neutrophil count. No significant differences were observed when it came to comorbidities at baseline. Thirty-two (86.5%) patients in the severe group and four (6.1%) in the mild group received corticosteroid treatment (p < 0.001). Most of these patients, 28 in the severe and two in the mild group, received the first dose before the first sampling timepoint.

**Table 1.**
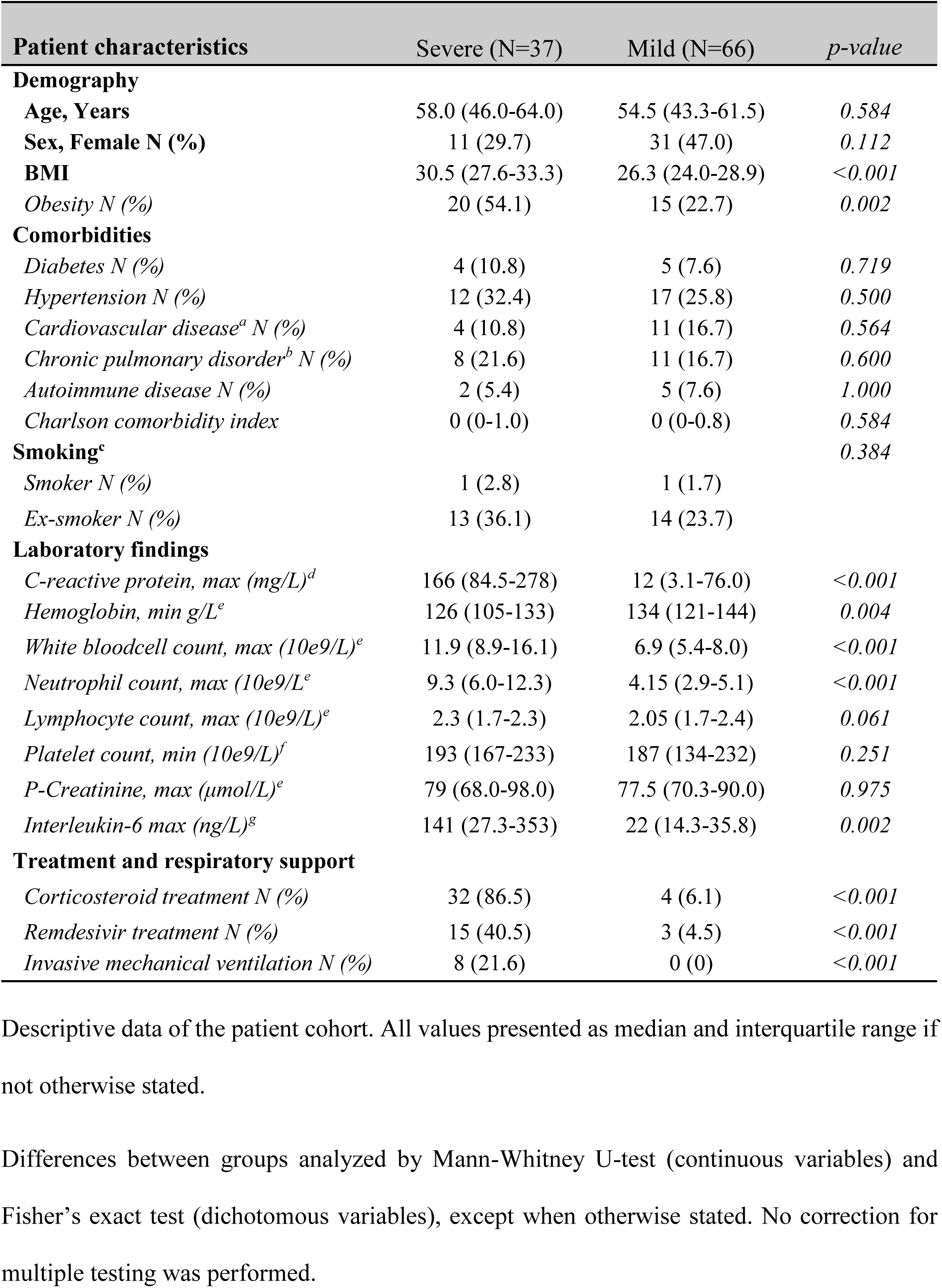
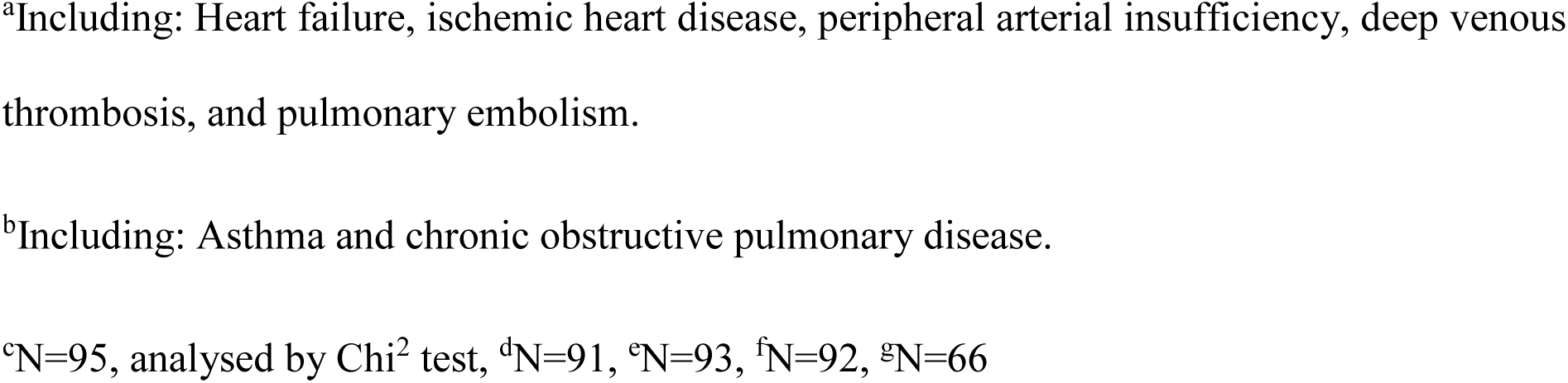
Demography and characteristics of COVID-19 patients.

The concentration of HA in plasma samples was determined using ELISA in order to assess the association between HA and disease severity. A general increase in HA concentrations was observed in samples taken during the acute phase of the disease in both patients with mild and severe COVID-19, as compared to healthy controls (figure 3A). The HA concentrations were even further increased in patients with severe COVID-19. Interestingly, although HA concentrations declined in the convalescent phase (mild p ˂ 0.0001, severe p ˂ 0.0001), they remained elevated compared to healthy controls, especially in patients with severe disease. A sex comparison of HA plasma concentrations in the acute phase showed a similar increase in HA in severe COVID-19 in women and men (figure 3B). However, the HA levels remained higher among women compared to men (p=0.043, data not shown) in the convalescent phase of mild COVID-19. Analyses assessing the impact of age showed an increase in HA concentrations in severe compared to mild COVID-19 in both older (60-89 years) and younger (18-59 years) patients, with a general trend towards higher HA concentrations in the older age group (mild disease: p=0.01, severe disease: p=0.17) (figure 3C). As we have seen that HA accumulated in the lungs during COVID-19, and systemic levels of HA remains high even after 12 weeks, we next wanted to investigate how the systemic HA levels would correlate to actual lung function. HA levels of acutely infected patients were associated with the percentage of predicted diffusion capacity (DL_CO_%pred) three to six months after infection (figure 3D-G). Lung function measurements were available in 70 out of 103 patients. The analysis was adjusted for sex, chronic lung disease, cardiovascular disease, hypertension, diabetes, smoking (current or previous), obesity (BMI ≥30), severity of COVID-19 (mild or severe), and age (20-59 and 60-89 years). Our results showed a negative correlation between HA levels in both the convalescent phase and acute phase with DL_CO_%pred (supplementary table 1 and 2).

**Figure 3.**
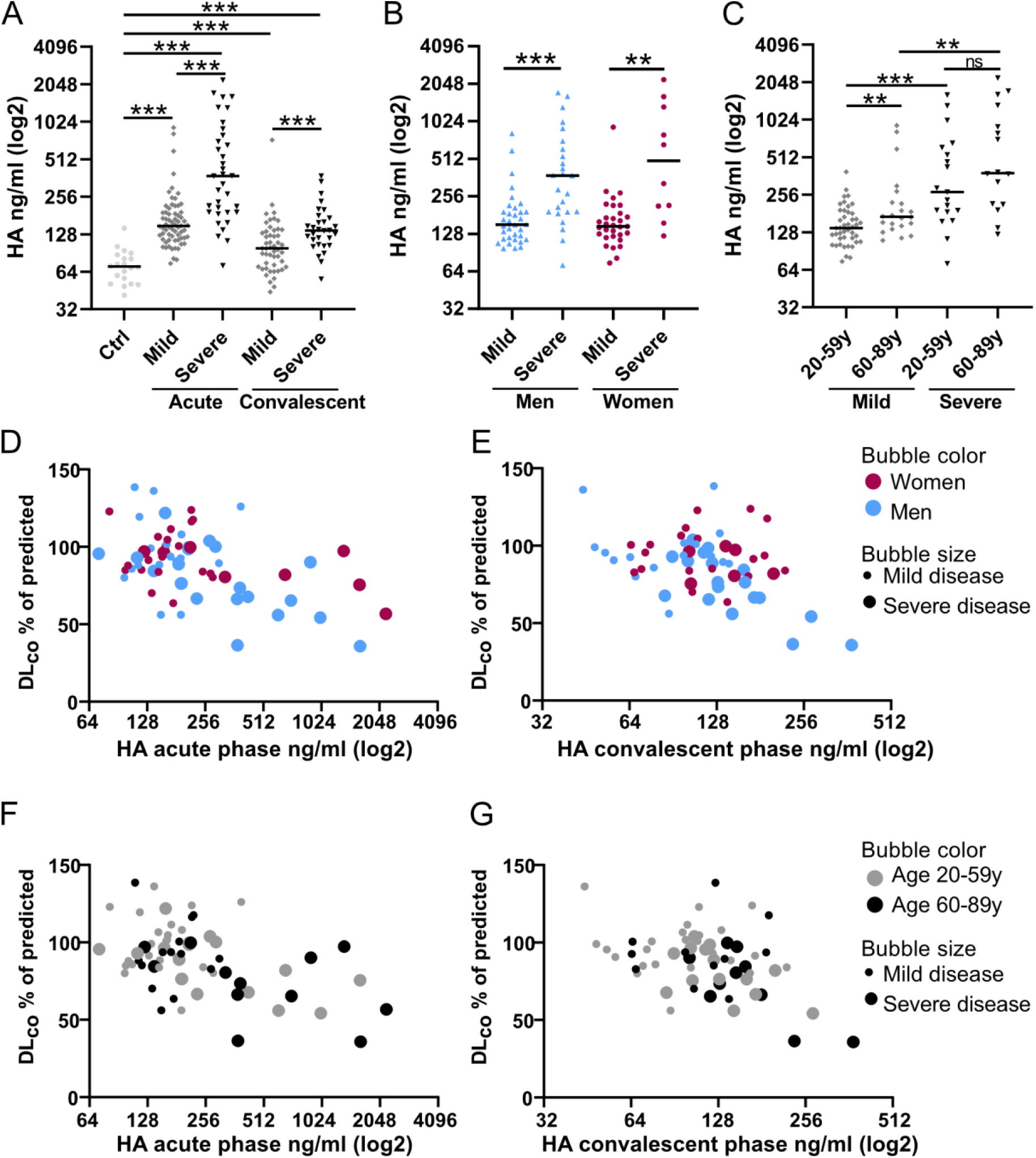
High concentrations of hyaluronan (HA) in plasma are associated with COVID-19 severity and long-term lung impairment. A) HA concentrations in plasma from patients with severe and mild COVID-19 compared to controls (Ctrl). Samples were taken during the acute phase (0-4 weeks from disease onset) and again during the convalescent phase of the disease (≥12 weeks). HA concentrations were determined by ELISA. B-C) HA concentrations in plasma during the acute phase grouped based on severity and B) sex or C) age. Each dot represents one patient, and the line represents the median. Severity was based on WHO Clinical Progression Scale (WHO-CPS) with patients requiring high-flow nasal oxygen treatment and/or admission to the intensive care unit during the acute phase of illness classified as “severe”, corresponding to WHO-CPS 6-9, and all other patients as “mild”, WHO-CPS 1-5. Statistical significance was calculated by Mann-Whitney U-test (*p < 0.05, **p < 0.01, ***p < 0.001). D-E) The percentage of predicted values of diffusion capacity (DL_CO_) related to HA concentrations in plasma in D) the acute phase or E) the convalescent phase divided by sex and disease severity. F-G) The percentage of predicted values of DL_CO_ related to HA concentrations in plasma in D) the acute phase or E) the convalescent phase divided by age and disease severity.

In summary, plasma concentrations of HA were substantially increased in COVID-19 patients and correlated to disease severity and reduced diffusion capacity. We then performed infection experiments in a human 3D-lung model to further study the potential mechanisms behind the increase of HA in COVID-19.

### SARS-CoV-2 infection causes an inflammatory response, which is counteracted by corticosteroids in a human 3D-lung model

We used a lung model that closely resembles the human respiratory tract based on primary human bronchial epithelial cells (HBECs) isolated from human donors. This model was used to characterize the cellular pathways affected by SARS-CoV-2 infection and the effects of corticosteroid treatment with respect to inflammation and HA metabolism. The cells were differentiated at an air-liquid interface (ALI) to form a polarized epithelium containing an apical layer of fully functional secretory and ciliated cells and an underlying layer of basal cells (18). The lung cultures were treated with, or without, the corticosteroid betamethasone, and infected with SARS-CoV-2 (figure 4A). The course of infection was monitored daily by collection of apical secretions containing released progeny virus and the viral load was quantified by qPCR. A distinct increase in viral RNA was observed over time, indicating an active viral replication in the HBEC ALI-cultures (figure 4B). Betamethasone-treated cultures showed reduced levels of released progeny virus. The infected HBEC ALI-cultures were harvested at 96h post infection and subjected to total RNA sequencing, to identify the mechanisms and pathways regulated by SARS-CoV-2 infection and betamethasone treatment. Statistical comparison of genes expressed in uninfected vs. infected HBEC ALI-cultures (figure 4C, supplementary table 3) identified 167 differentially expressed genes (DEGs) that were upregulated upon SARS-CoV-2 infection, with a strong enrichment of genes involved in the immune response, especially type I interferon (IFN-I) signaling. A similar statistical comparison of infected HBEC ALI-cultures with or without betamethasone treatment was performed to evaluate the effect of betamethasone treatment. This identified 102 genes that were upregulated, and 229 genes that were downregulated in the betamethasone-treated cultures (figure 4D, supplementary table 3). Interestingly, 73 of the 167 genes upregulated by infection (figure 4C) were downregulated by betamethasone treatment (figure 4D). A heatmap displaying the mean expression values of the individual overlapping genes showed a clear upregulation of these genes by SARS-CoV-2 infection, which was counteracted by betamethasone treatment (figure 4E). Several of the affected genes were involved in inflammatory responses, correlating well with the anti-inflammatory activity of betamethasone. We also collected basolateral samples from the HBEC ALI-cultures at 96h post infection and analyzed these with a targeted cytokine panel based on proximity extension assay, allowing a quantitative concentration measurement of each cytokine (figure 4F). Of the ten cytokines that showed significant changes, eight were downregulated by betamethasone treatment, but not affected by the infection. CXCL10 and CXCL11 were both upregulated by the SARS-CoV-2 infection and subsequently downregulated by steroid treatment.

**Figure 4.**
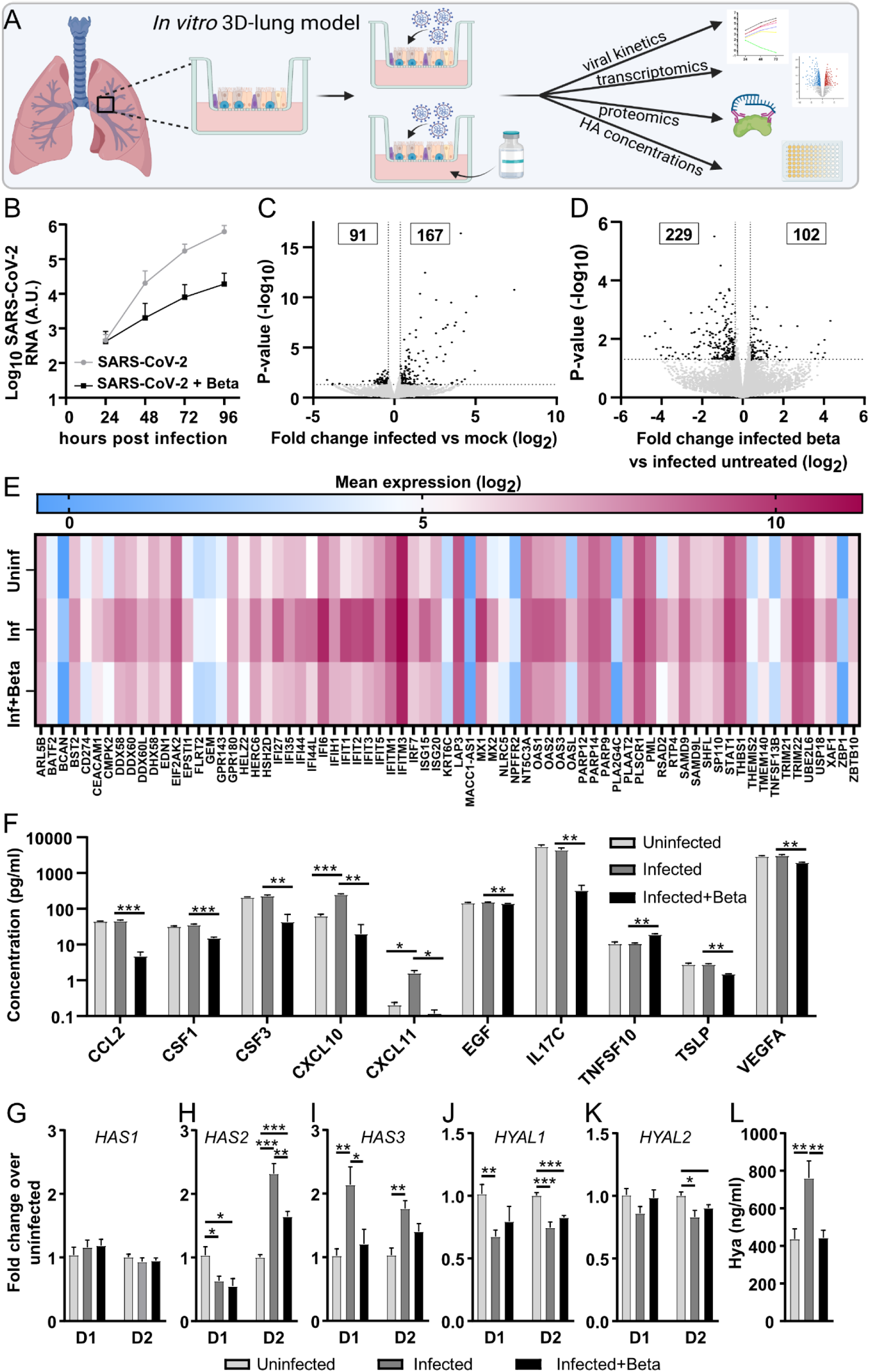
Inflammatory response and effect of corticosteroid treatment in a SARS-CoV-2 infected 3D-lung model. A) Schematic overview of the primary 3D-lung model that was infected with SARS-CoV-2 in the presence or absence of corticosteroids to investigate the effect on inflammation and hyaluronan metabolism. B) The lung cultures based on differentiated primary human bronchial epithelial cells at an air-liquid interface were pretreated with/without betamethasone (Beta) in the basal media for 20 h prior to infection with SARS-CoV-2 (multiplicity of infection=0.5). The accumulated viral release from the apical side of the cultures was quantified by qPCR at indicated timepoints. C-D) Volcano plot showing differentially expressed genes between C) infected vs uninfected (mock) lung cultures and D) infected lung cultures with betamethasone vs without betamethasone treatment. The statistical *p*-value (-log_10_) is plotted against the fold change in gene expression (log_2_). Dotted lines highlight the significance cut off corresponding to a fold change of 1.3 and p-value=0.05. E) Heatmap displaying the mean expression (log_2_) of the overlapping genes in uninfected, infected and infected + betamethasone-treated lung cultures. F) Cytokine levels in basolateral samples from HBEC ALI-cultures collected at 96 h post infection analyzed by Proximity Extension Assay (Olink). The gene expression levels of HA synthases G) *HAS1*, H) *HAS2* and I) *HAS3* along with the hyaluronidases J) *HYAL1* and K) *HYAL2* were determined by qPCR 120 h post SARS-CoV-2 infection of lung cultures from two different donors (D1 and D2). L) HA concentrations were determined by ELISA in apical secretions from lung cultures 120 h post infection. Mean values and SEM are shown, statistical significance was calculated by unpaired t-test (\**p* < 0.05, \*\**p* < 0.01\*\*\**p* < 0.001).

### SARS-CoV-2 affects HA metabolism by upregulation of HA synthases and downregulation of hyaluronidases

To further investigate the direct effect of SARS-CoV-2 on HA synthesis and degradation, expression levels of the three human hyaluronan synthases, *HAS1, HAS2* and *HAS3* and the two major hyaluronidases, *HYAL1* and *HYAL2*, were determined in HBEC ALI-cultures from two different donors five days post infection. No effect was seen on *HAS1* expression, but SARS-CoV-2 infection increased *HAS*2 in donor 2 and *HAS3* expression in both donors (figure 4G-I). Betamethasone treatment of infected cultures resulted in a reduction of the upregulated *HAS2* expression in donor 2 and *HAS3* in donor 1 (figure 4H and I). Additionally, SARS-CoV-2 infection decreased expression of hyaluronidase *HYAL1* in both donors and *HYAL2* in donor 2 (figure 4J and K). The hyaluronidase genes were not significantly affected by betamethasone treatment (figure 4J and K). Both upregulation of HA synthases and downregulation of degrading hyaluronidases can induce an increase in HA concentrations. Indeed, measurements of HA concentrations in the apical secretions from HBEC ALI-cultures confirmed the transcriptional changes with an increase in HA in cultures infected by SARS-CoV-2, which were kept at baseline concentrations when simultaneously treated with betamethasone (figure 4L).

Taken together, our results identified genes involved in viral defense and inflammation affected by SARS-CoV-2 infection. Betamethasone treatment demonstrated a general counteraction against the viral transcriptional effect. SARS-CoV-2 infection and betamethasone treatment also affected genes with potential impact on HA production and degradation. In addition to the effects on HA synthases and hyaluronidases, we observed an upregulation of transcription factors early growth response 1 and 2 (*EGR1, 2*) upon SARS-CoV-2 infection and a downregulation of lactate dehydrogenase A (*LDHA*) and TP53 induced glycolysis regulatory phosphatase (*TIGAR*) after betamethasone treatment, all of which are involved in the glycolysis and therefore affect HA metabolism (supplementary table 3). Based on our results, we present a model where the positive action of betamethasone in severe COVID-19 patients is a combined action of a reduced inflammatory response (figure 5A, supplementary table 3) and a reduction of the pathological overproduction of HA upon SARS-CoV-2 infection (figure 5B, supplementary table 3).

**Figure 5.**
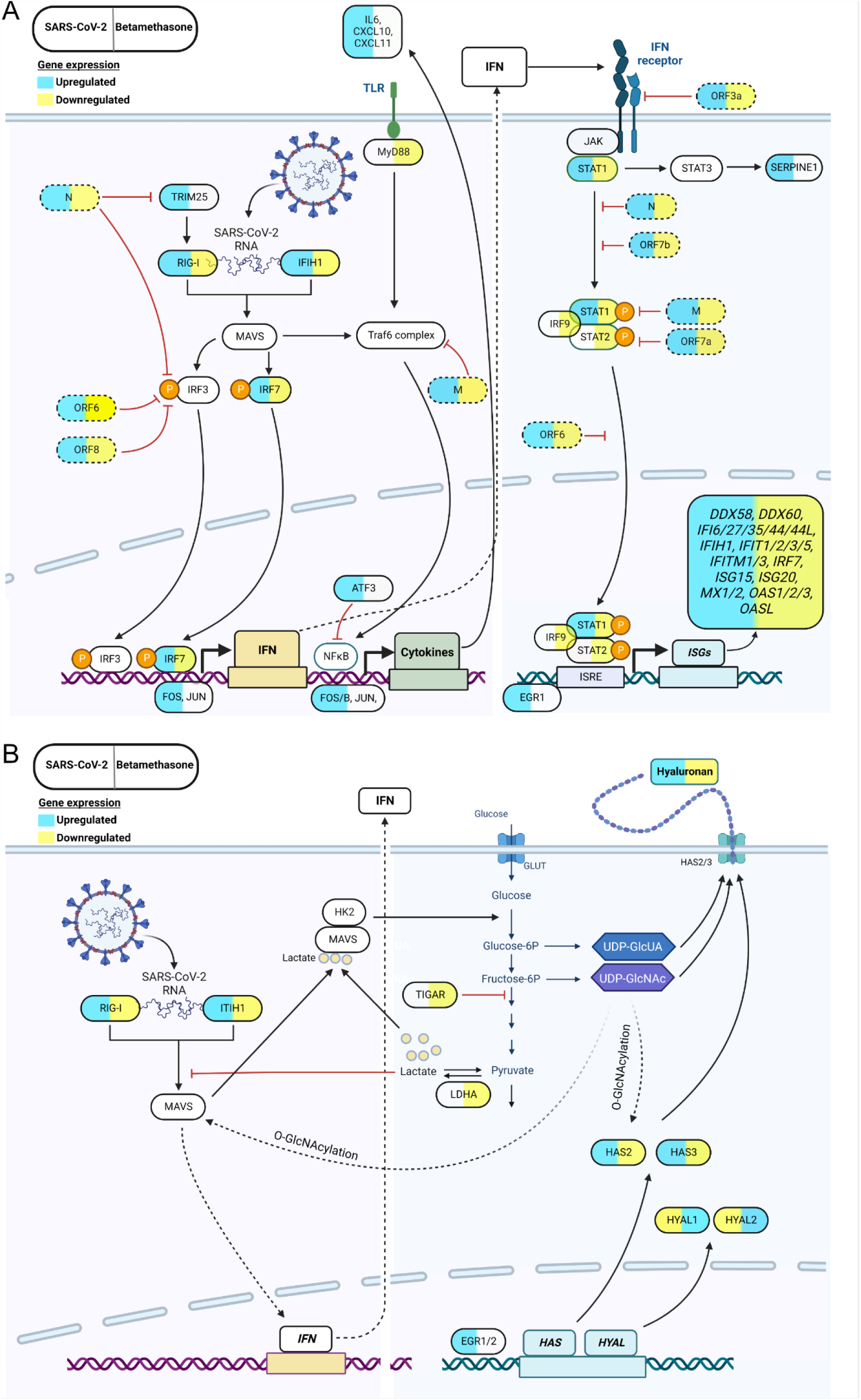
The effect of SARS-CoV-2 and betamethasone on cellular pathways regulating IFN-I responses and hyaluronan (HA) production in a human *in vitro* 3D-lung model. Schematic presentation of a cell infected with SARS-CoV-2 with or without betamethasone treatment and affected cellular pathways regulating A) the IFN-I response and B) HA production. The left side of the gene shows the effect on gene expression upon SARS-CoV-2 infection, and the right side shows effect of betamethasone treatment in infected lung cultures. Blue color corresponds to an increase in gene expression and yellow indicates a decrease in gene expression. Viral genes are represented by dotted lines.

## DISCUSSION

We have previously shown that lungs from deceased COVID-19 patients are filled with a clear liquid jelly consisting of HA, which impairs the capillary-alveolar gas exchange and leads to respiratory failure. In this study, we investigated the morphology of the lung in 3D, the presence of fragmented HA during severe COVID-19 and the correlation between systemic HA levels and diffusion capacity of the lungs after recovery. We also show the mechanisms behind the pathological increase in HA levels, which were counteracted by corticosteroid treatment, to possibly identify new therapeutic targets using a human *in vitro* 3D-lung model.

Morphological studies of tissue are commonly done by immunohistochemistry of thin tissue slices. These have the advantage of a high resolution down to subcellular level, and many cellular markers can be used to identify different features. However, it is much more difficult to analyze the composition and morphology of lung tissue especially the alveolar volume or surface areas. We therefore used LSFM to study the morphology of lung necropsies and biopsies in 3D. Our results showed a reduction in the number and size of the alveoli, as well as the surface area used for gas exchange at end stage of COVID-19 compared to healthy controls. Interestingly, LSFM has been used previously in a ferret model of COVID-19, but no similar obstructions were seen in their lungs (19). With histochemistry we showed that the alveoli were filled with HA and this HA was highly fragmented in the COVID-19 necropsies, likely contributing to the inflammatory milieu visualized by large infiltrates of neutrophils. The HA was also fragmented in severe COVID-19 in both nasopharyngeal and endotracheal aspirates. Interestingly, hyper-induction of HA in the lungs appears in many respects to be a reversible process. Even though reduced diffusion capacity correlates to disease severity (15) and is related to severity of radiologic lung involvement at admission (20), there is still restitution. This is supported by our data where the recovered COVID-19 patient did not present with a reduced mean alveolar surface area compared to the healthy controls. However, the presence of collagen coils that disrupt alveolar walls in necropsy COVID-19 lungs suggests that the filling of HA in the alveoli precedes the development of collagen fibrosis and irreversible lung damage. This is supported by previous findings in both human inflammatory diseases and in a lung damage rat model (6, 21, 22). The role of HA size in fibrosis is not clear, but it has been demonstrated that the interaction of fragmented HA and the receptor HA-mediated motility receptor (RHAMM), increased the infiltration of fibroblasts, leading to uncontrolled wound healing with collagen production and fibrosis (23).

We showed that despite an overall reduction in systemic HA concentrations during the convalescent phase (≥12 weeks), the plasma levels did not normalize in either mild or severe COVID-19 patients and remained significantly higher compared to the control group. Prior studies have demonstrated an elevation of HA concentration associated with disease severity in the acute phase of COVID-19 (10, 11), but the long-term consequences of COVID-19 on HA levels have not previously been thoroughly investigated. The tissue half-life of HA ranges from half a day to two-three days, and the half-time in blood is even shorter (24). The sustained elevation of plasma HA concentrations demonstrated here therefore suggests an imbalance in the production and degradation of HA that remains for at least 12 weeks after disease onset, even after mild disease. The correlation of high plasma levels of HA in both acute and convalescent phase with reduced diffusion capacity also implies that plasma HA may be used as a predictive biomarker for future lung function impairment.

The molecular mechanisms behind the observed increase in HA synthesis upon SARS-CoV-2 infection remain to be clarified. Based on the results of this study, we here propose a model in which SARS-CoV-2 infection causes transcriptional changes of genes involved in HA metabolism, which are partially counteracted by corticosteroid treatment. In our human *in vitro* 3D-lung model, SARS-CoV-2 infection caused an upregulation of the transcription factors *EGR1* and *EGR2*, which in turn activated the expression of HA synthases *HAS2* and *HAS3.* At the same time, SARS-CoV-2 infection decreased the degrading hyaluronidases. However, it is possible that additional pathways can affect HA production. It was recently shown that specific RNA sequences in the SARS-CoV-2 genome can activate expression of *HAS2* and, consequently, increase HA synthesis (25). In addition, changes to the glycolysis pathway may affect HA levels, due to an increased production of HA precursor molecules (26). Interestingly, others have shown that SARS-CoV-2 infection and subsequent replication induce an increased glucose metabolism in infected cells (27, 28). Corticosteroid treatment reduced the overall HA concentration in our 3D-lung model during SARS-CoV-2 infection. Interestingly, besides counteracting the SARS-CoV-2-induced effect on hyaluronan synthases, corticosteroid treatment of the infected 3D-lung model also decreased the expression of lactate dehydrogenase A (LDHA) and TIGAR, which may both contribute to decreased HA production via the glycolysis pathway (29, 30).

Today, one of the few evidence-based treatment options for patients with severe to critical COVID-19 is corticosteroids (12). However, the timing of treatment seems to be crucial. Early treatment of COVID-19 patients with corticosteroids has been shown to have less positive effect than treating severely ill patients (12). The increased levels of HA in the blood from both mild and severe cases of COVID-19, shown by us and others (10, 11), indicate a dysregulated HA metabolism in COVID-19, which was also supported by the findings in our 3D-lung model. Hyaluronan production and/or degradation therefore poses an attractive treatment target for severe COVID-19. Hymecromone, an FDA-approved drug for treatment of biliary spasm, has a more direct action against HA synthesis and could be used to specifically treat the overproduction of HA. Such a high-precision treatment could avoid some of the negative effects of corticosteroids and enable treatment at an earlier stage to prevent disease progression. A recent clinical trial with hymecromone showed efficient inhibition of COVID-19 progression and warrants further investigations (31).

In summary, our results showed destructed lung morphology with markedly decreased gas exchange surface in COVID-19, fragmented inflammatory HA in both nasopharyngeal aspirate and endotracheal aspirate and sustained increased levels of HA in peripheral blood in COVID-19 patients. The increased HA expression could be explained by the mechanisms identified in our *in vitro* lung model including an imbalance in HA production and degradation induced upon SARS-CoV-2 infection. We also show that elevated systemic HA levels both in acute COVID-19 and convalescence phase negatively correlate with diffusion capacity in the lungs during convalescence. Further studies are needed to evaluate systemic HA as a biomarker for long term lung function impairment. We show that HA is an important factor in COVID-19 pathogenesis, and studies on targeting HA metabolism as an alternative or complementary treatment to corticosteroids to reduce the acute and long-term health consequences of COVID-19 are warranted.

## METHODS

### Tissue pre-processing for optical 3D-imaging

Lung biopsies from four healthy controls, three deceased COVID-19 patients and one patient that recovered from COVID-19 were washed in PBS, fixed in formalin overnight, dehydrated successively into methanol (MeOH) (25%, 50%, 75%, 100%, 15 min each step) and stored at - 20°C until use. Prior to clearing, the lung samples were treated in bleaching solution (H_2_O_2_: dimethyl sulfoxide: MeOH, 3:1:2) overnight at room temperature (RT). The bleaching solution was refreshed, and the samples were incubated for an additional 6 hours at RT. Following bleaching, the samples were rehydrated into PBS (25%, 50%, 75%, 100%, 15 min each step) and mounted in 1.5% low-melting point agarose (Lonza™ SeaPlaque™ Agarose, Lonza, USA). After solidifying overnight, the agarose pieces were cut into cuboids and dehydrated into MeOH (25%, 50%, 75%, 100%, 15 min each step). The samples were subsequently washed in MeOH for 2x24 hours on rotation in order to remove any residual water. Following MeOH washes, the samples were optically cleared in BABB (benzyl alcohol, benzyl benzoate, 1:2) changing solution every 12 hours until the biopsies were completely cleared.

### Light sheet fluorescence microscopy and 3D image processing

High resolution 3D images of the lungs were acquired using a UltraMicroscope II (Miltenyi Biotec, Germany) fitted with an 1x Olympus objective (Olympus PLAPO 2XC) and a lens corrected dipping cap MVPLAPO 2x DC DBE objective attached to an Olympus MVX10 zoom body with a 3000 step chromatic correction motor. The lung regions of interest were captured at a magnification of 1.6x with a scan depth of 1000 µm, a dynamic focus range across the specimen capturing 10 images per section, and a step-size of 5 µm yielding a voxel size of 1.89 x 1.89 x 5 µm. The samples were scanned for autofluorescence with the filters: Ex 470/40, Em: 525/50 using an exposure time of 300ms. Optical sections were saved in *ome.tif format native to the ImspectorPro software (version 7.0.124.9 LaVision Biotex GmbH, Germany). The *ome.tif files were converted into 3D projection *ims files using the Imaris file converter (version 9.9.1, Bitplane, UK) 3D volumes were generated using the built in surfacing method of Imaris (version 9.8.0, Bitplane, UK). To quantify the “empty space volume”, roughly translating to the alveolar volume, the threshold was set to only include hypointense space using absolute intensity for surfacing, with a surface grain size set to 0.8 µm. Further, an exclusion filter removing objects consisting of less than 10 voxels was applied to remove general noise. The anatomy surface was generated and set to include all signal above the hypointense signal with a surface grain size of 3.78 µm and a 10 voxel exclusion filter. Surfaces generated from hypointense regions outside the tissue volume were manually excluded when possible. Volumes and area of the segmented surfaces were extracted from Imaris as Excel (Microsoft, Office 365, version 2301) *.XML file format for quantification.

### Isolation and fragmentation of HA from lung biopsies and aspirates

#### (i) HA isolation

Cellular secrets were dried in a Savant SpeedVac DNA 110 vacuum concentrator (Thermo Fisher Scientific, MA, USA). Proteins and nucleic acids were digested with proteinase K (Sigma-Aldrich, St Louis, MO, USA) and benzonase nuclease (Sigma-Aldrich) on two consecutive days. At the end of each digestion, chloroform was added to each sample and the extracted aqueous phase was solvent exchanged to 0.1 M NaCl using Amicon Ultra 3K concentration units (Millipore, Billerica, MA, USA) followed by overnight precipitation in 99% ethanol. Sulphated glycosaminoglycans and remaining non-HA contaminants were removed with anion-exchange mini spin columns (Thermo Fisher Scientific, MA, USA), based on NaCl binding. Finally, to remove salt the sample was solvent exchanged to 20 mM ammonium acetate (pH 8.0) in Amicon Ultra 3K concentration units.

#### (ii) HA molecular mass analysis

HA mass analysis was undertaken using a gas-phase electrophoretic mobility molecular analysis (GEMMA) (TSI Corp., MN, USA). The molecule diameter analyzed in the GEMMA was converted to molecular mass by analyzing HA standards ranging from 30 kDa to 2500 kDa (Hyalose, OK, USA). The area under the curve corresponds to the number of molecules in the GEMMA analysis.

### Study design and study population

Data and clinical samples were obtained from the CoVUm study, a prospective, multicenter observational study of COVID-19 including patients from Umeå and Örebro, and coordinated from Umeå University, Sweden (www.clinicaltrials.gov identifier NCT 04368013). The study protocol and cohort have been described in detail earlier (32). Non-hospitalized patients aged ≥15 years and hospitalized patients aged ≥18 years with a positive PCR-test for SARS-CoV-2 were enrolled in the study. Written informed consent was obtained from all participants or their next of kin before the first sampling timepoint. At data export on May 4, 2022, a total of 543 participants, enrolled between 27 April 2020 and 28 May 2021, had been registered in the CoVUm database. Seven of the participants were excluded since they did not fulfill the inclusion criteria (false positive PCR-tests for SARS-CoV-2). Out of the remaining 536 participants, all patients classified as severely ill during the acute phase of the disease, and with available blood samples, were selected for this study. In addition, 66 patients with mild COVID-19 were selected from the remaining study cohort. Participants were classified as “severe” if they required high-flow nasal oxygen treatment (HFNO) and/or was admitted to the intensive care unit (ICU) during the acute phase of illness, corresponding to WHO Clinical Progression Scale (WHO-CPS) 6-10 (17). All other participants were classified as “mild”, corresponding to WHO-CPS 1-5. Blood samples were obtained at acute phase (0-4 weeks after onset) and convalescent phase (≥12 weeks). For study outline, see figure 2A. A healthy control group, consisting of plasma samples collected from anonymous blood donors was included for reference. The study was performed according to the Declaration of Helsinki and approved by the Swedish Ethical Review Authority.

### Data collection and clinical samples

All clinical metadata including age, sex, Charlson Comorbidity Index (CCI) (33), tobacco use, medication at enrollment, body mass index, type of respiratory support and medical treatment were collected and managed using the REDCap electronic data capture tools hosted at Umeå University (34, 35). Clinical chemistry data of conventional inflammatory markers were extracted retrospectively from the patients’ electronic medical records. The highest and lowest values of each biomarker from each individual, during the first 180 days after symptom onset were extracted for further analysis. Plasma samples used for HA analysis were collected at each timepoint in 6 ml EDTA tubes (BD Diagnostics).

### Measurement of hyaluronan concentration in plasma samples

Plasma hyaluronan (HA) concentrations were measured with a competitive HA-binding protein-based enzyme-linked immunosorbent assay (ELISA)-like concentration measurement kit (K-1200; Echelon Biosciences Inc., Salt Lake City, UT, USA), according to the manufacturer’s instructions. Samples were run in duplicate and a coefficient of variation (CV) < 10% was considered acceptable. Absorbance was measured on a ThermoMultiskan Ascent (Thermo Fisher Scientific, MA, USA) and plotted by polynomial regression against the concentration of the standard curve.

### Lung function tests

Lung function tests were conducted 3-6 months after study enrolment for non-hospitalized patients, or discharge from hospital for patients hospitalized during the acute phase of illness, as previously described (15). Reference values for DL_CO_ were calculated using The Global Lung Function Initiative (GLI) Network guidelines.

### Assessment of hyaluronan metabolic pathways upon SARS-CoV-2 infection in an *in vitro* 3D-lung model

#### (i) Generation of the human primary 3D-lung model

Primary human bronchial epithelial cells (HBEC) were isolated from proximal airway tissue obtained with informed consent from two patients, who underwent thoracic surgery at the University Hospital, Umeå, Sweden (ethical permission approved by the Regional Swedish Ethical Review Authority in Umeå). HBECs were grown and differentiated at an air-liquid interface (ALI) forming an *in vitro* 3D-lung model as previously described (16). In short, HBECs were grown and differentiated on 6.5 mm semipermeable transwell inserts (0.4 µm Pore Polyester Membrane Insert, Corning) and after two weeks at air-liquid interface (ALI) the cultures reached full differentiation, which was assessed using light microscopy focusing on epithelial morphology, presence of ciliated cells, and mucus production along with immunofluorescence staining for ciliated cells (acetylated-tubulin, T6793, Sigma) and goblet cells (muc5AC, Ab-1 (45M1), #MS-145-P, ThermoFisher).

#### (ii) Betamethasone treatment

Fully differentiated HBEC-ALI cultures were either mock treated or treated with betamethasone (Alfasigma) 20 h prior to infection by addition of 700 µl fresh basal media containing 0.3 µM betamethasone. For study outline see figure 4A. Shortly before infection, the basal media was exchanged once more and betamethasone was replenished in the treated wells. Betamethasone was replenished every 24 h post infection.

#### (iii) SARS-CoV-2 infection

The clinical isolate SARS-CoV-2/01/human/2020/SWE (GeneBank accession no. MT093571.1) was kindly provided by the Public Health Agency of Sweden. Vero E6 cells were cultured in Dulbecco’s modified Eagle’s medium (DMEM, D5648 Sigma) supplemented with 5% FBS (HyClone), 100 U/mL penicillin and 100 µg/ml streptomycin (PeSt, HyClone) at 37°C in 5% CO_2_. Propagation of the virus was done once in Vero E6 cells for 72 h and titration was done by plaque assay. The apical side of the HBEC ALI-cultures were rinsed three times with warm PBS shortly before infection. 1.5x10^4^ plaque forming units (PFU) of SARS-CoV-2 was added to the apical compartment in a total volume of 100 µl infection medium (DMEM / PeSt), corresponding to an approximate multiplicity of infection (MOI) of 0.05. The HBEC ALI-cultures were incubated at 37°C and 5% CO_2_ for 2.5 h before the inoculum was removed and the cultures were washed with PBS to remove residual medium.

#### (iv) Sample collection

Accumulated progeny virus and secretions were collected from the apical side of the HBEC ALI-cultures every 24 h by addition of 100 µl warm PBS to the apical chamber followed by a 1 h incubation at 37°C and 5% CO_2_. The collected samples were stored at –80°C until RNA extraction. The progression of the infection was monitored for four days (96 h post infection).

#### (iv) Virus quantification by qPCR

Viral RNA secreted from HBEC ALI-cultures was extracted from 50 µl of the apical samples using the QIAmp Viral RNA kit (Qiagen) following the manufacturer’s instructions, and cDNA was synthesized from 10 µl of eluted RNA. RT-qPCR for SARS-CoV-2 RNA was performed in duplicates on a StepOnePlus^TM^ Real-Time PCR System (Applied Biosystems) using the qPCRBIO Probe Mix Hi-ROX (PCR biosystems) and primers (Forward: GTCATGTGTGGCGGTTCACT, Reverse: CAACACTATTAGCATAAGCAGTTGT) and probe (CAGGTGGAACCTCATCAGGAGATGC) specific for viral RdRp.

### Transcriptomics, total RNA sequencing

At 96 h post infection the mock treated (n=3), infected (n=3) and infected + betamethasone treated (n=3) HBEC ALI-cultures were washed three times on both sides with PBS. The HBEC ALI-cultures were then lysed and RNA extraction was done using the NucleoSpin RNA II kit (Macherey-Nagel) following the manufacturer’s instructions. RNA-seq libraries were prepared using the Smart-seq2 method, and sequenced on an Illumina NextSeq 500 (75PE, v2.5 High Output kit). STAR 2.7.1a was used to align the reads against a reference genome consisting of GRCh38 and Sars_cov_2.ASM985889v3. A gene expression table was produced using featureCounts (36). Differential expression analysis was performed using DESeq2 (37).

### Cytokine quantification

At 96 h post infection, samples were collected from the basal chambers of each of the HBEC ALI-cultures. The samples were inactivated with Triton X-100 at a final concentration of 1% followed by incubation at room temperature for 3 h. The levels of 45 cytokines were quantified by Proximity Extension Assay (Olink Target 48 Cytokine panel at Affinity Proteomics Uppsala, SciLifeLab Sweden), which gives absolute (pg/mL) and relative (normalized protein expression, NPX) concentration measurements of 45 pre-selected cytokines.

### Quantification of HA synthases and hyaluronidases by qPCR

Total RNA from cells were extracted using the Nucleo-Spin RNA II kit (Macherey-Nagel). 1000 ng of RNA was used as input for cDNA synthesis using High-capacity cDNA Reverse Transcription kit (Thermo Fisher). Cellular HA synthases (*HAS1, 2* and *3*) and hyaluronidases (*HYAL1* and *2*) were quantified using qPCRBIO SyGreen mix Hi-ROX (PCR Biosystems) and QuantiTect primer assay (Qiagen, *HAS1*;QT02588509, *HAS2*;QT00027510, *HAS3*;QT00014903, *HYAL1*;QT01673413, *HYAL2*;QT00013363) with actin as a housekeeping gene (QT01680476) and run on a StepOnePlus^TM^ Real-Time PCR System (Applied Biosystems).

### Measurement of HA concentration in apical secretions from HBEC ALI-cultures

Apical secretions from HBEC ALI-cultures were collected at 120 h post infection and HA concentrations were measured with a Hyaluronic Acid AlphaScreen Assay (K-5800; Echelon Biosciences Inc., Salt Lake City, UT, USA), according to the manufacturer’s instructions. AlphaScreen beads from the Histidine (Nickel Chelate) Detection Kit (PerkinElmer, MA, USA) was used. Chemiluminescent emission was measured on a SpectraMax i3x (Molecular Devices, CA, USA) and plotted by polynomial regression against the concentration of the standard curve.

### Statistical analysis

Statistical analysis was performed with Graphpad Prism 9 and Jamovi version 2.2.5 (The jamovi project (2021)). jamovi (Version 1.6). Retrieved from https://www.jamovi.org). Descriptive variables of the patient cohort were analyzed by Mann-Whitney U-test (continuous variables) and Fisher’s exact test or Chi2 test (dichotomous variables). Missing data were handled by complete case analysis. No correction for multiple testing was performed. Multiple linear regression was used to explore the association between DL_CO_ and HA levels in plasma during the acute illness and the convalescent phase.Independent variables were sex, chronic lung disease, cardiovascular disease, hypertension, diabetes, smoking (current or previous), obesity (BMI ≥30), severity of COVID-19 (mild or severe), and age (20-59 and 60-89 years). The log transformation was used to address skewed data in HA.

### Data availability

The CoVUm data cannot be made publicly available, according to Swedish data protection laws and the terms of ethical approval that were stipulated by the Ethical Review Authority of Sweden. Access to data from the CoVUm database is organized according to a strict data access procedure, to comply with Swedish law. For all types of access, a research proposal must be submitted to the corresponding authors for evaluation. After evaluation, data access is contingent on vetting by the Ethical Review Authority of Sweden, according to the Act (2003:460) concerning the Ethical Review of Research Involving Humans.

## Supporting information

Supplementary Movie 1. Healthy

Supplementary Movie 2. COVID-19

Supplementary Movie 3. Recovered

## Data Availability

All data produced in the present study are available upon reasonable request to the authors

## Acknowledgement

The authors thank the team at Affinity Proteomics Uppsala, SciLifeLab Sweden, for providing assistance with Olink assays. We acknowledge our study nurses Ida-Lisa Persson and Anna Kauppi at the Department of Infectious Diseases in Umeå and Christine Degner, Anna Segerås and Lena Irvhage in Örebro, and the personnel at the Clinical Research Center at Umeå University Hospital and Örebro University Hospital for enrolment and sampling of study participants. We also acknowledge Umeå Center for Microbial Research (UCMR); the Biochemical Imaging Center at Umeå University (BICU), and the National Microscopy Infrastructure for microscopy support (NMI; VR-RFI 2019-00217). Figure 1A, 2A, 4A and 5 were created using BioRender.com.

## SUPPLEMENTARY INFORMATION

**Supplementary figure 1.**
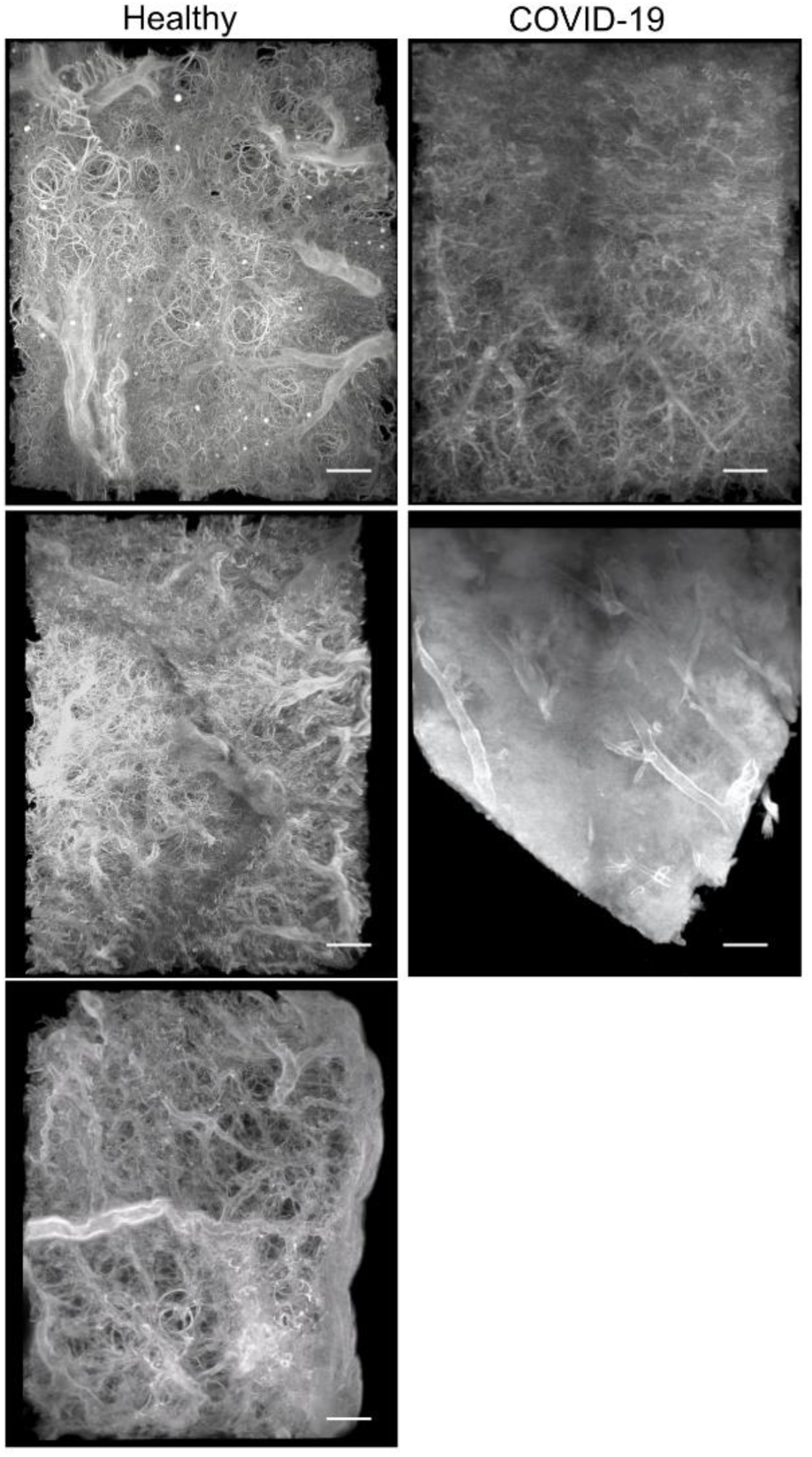
Light sheet fluorescent microscopy of biopsies from deceased COVID-19 patients and healthy donors. Maximum intensity projections of three healthy donors and two COVID-19 necropsies, scale bar is 500 µm.

**Supplementary figure 2.**
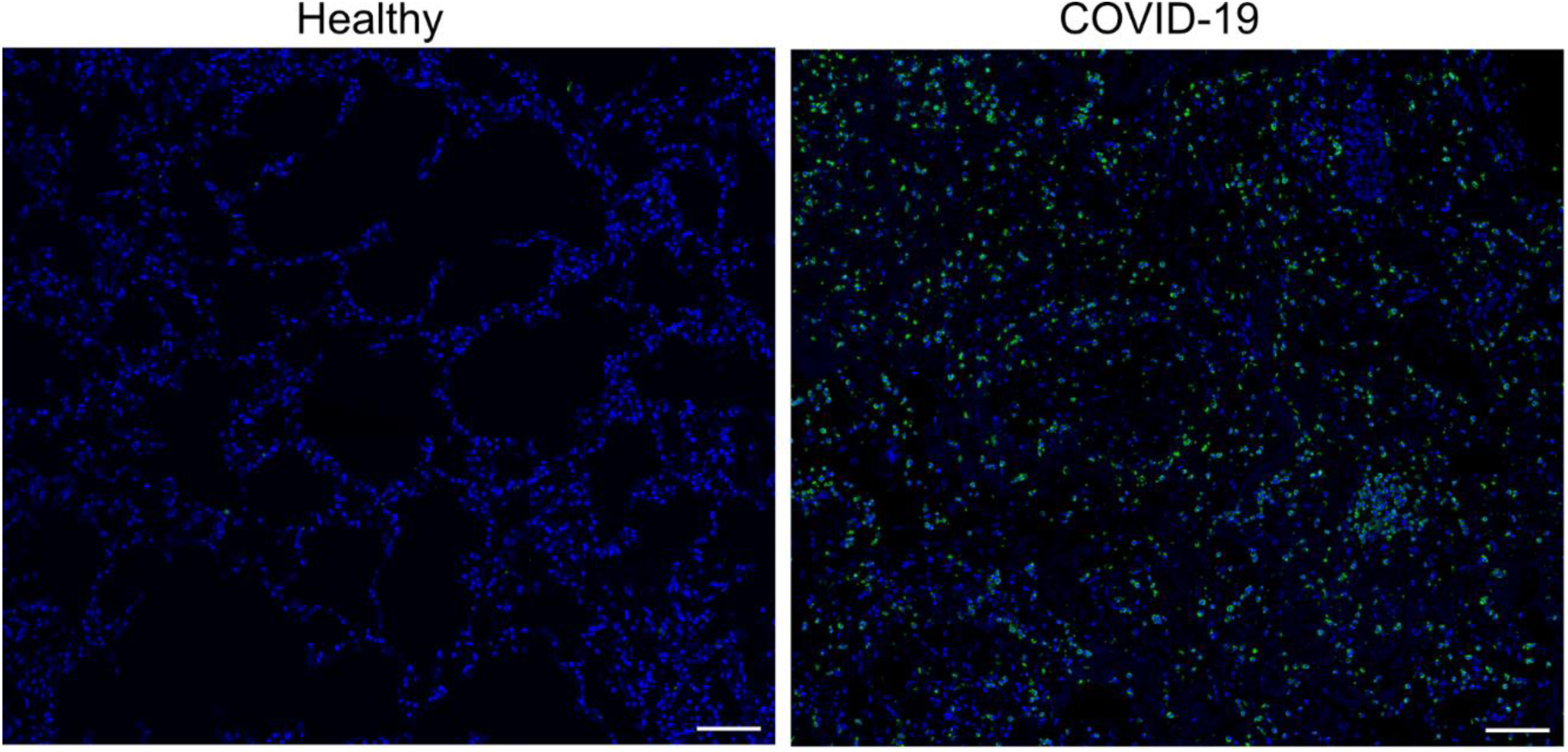
Neutrophil infiltration of COVID-19 lung necropsy. Confocal images on paraffin sections from healthy and COVID-19 lung biopsies stained with Dapi (blue) and neutrophil elastase (green). Representative pictures from imaged biopsies, scalebar is 100 µm.

**Supplementary Table 1.**
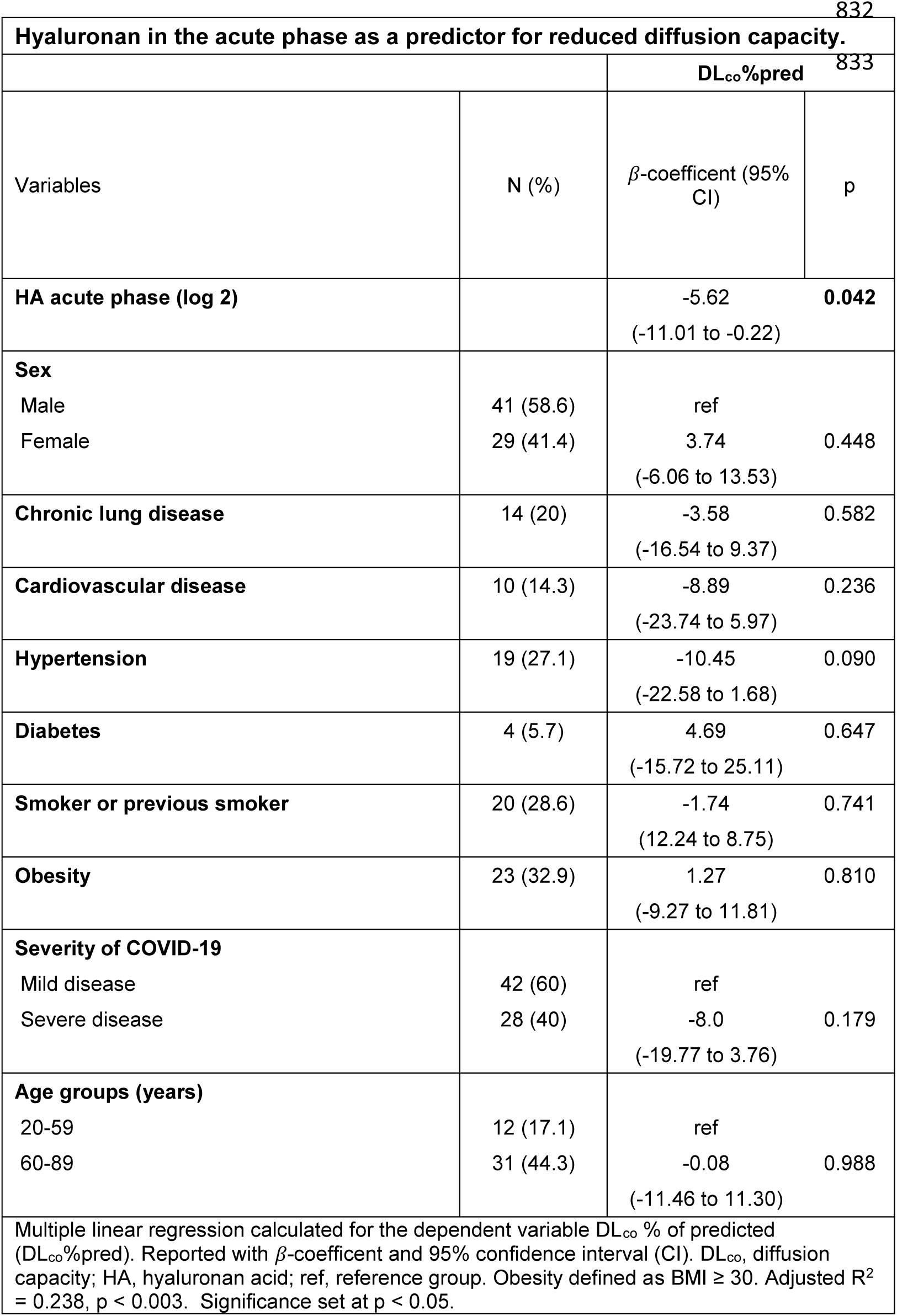

**Supplementary Table 2.**

| **Hyaluronan in the convalescent phase as a predictor for reduced diffusion capacity.** |  |  |  |
| --- | --- | --- | --- |
|  |  | **DL<sub>co</sub>%pred** |  |
| Variables | N (%) | β-coefficient (95% CI) | p |
| **HA convalescent phase (log 2)** |  | -9.51<br>(-18.21 to -0.82) | **0.033** |
| **Sex** |  |  |  |
| Male | 41 (58.6) | ref |  |
| Female | 29 (41.4) | 4.62<br>(-5.45 to 14.68) | 0.361 |
| **Chronic lung disease** | 14 (20) | 0.68<br>(-14.63 to 15.99) | 0.929 |
| **Cardiovascular disease** | 10 (14.3) | -17.64<br>(-33.48 to -1.80) | **0.030** |
| **Hypertension** | 19 (27.1) | -10.03<br>(-22.88 to 2.83) | 0.123 |
| **Diabetes** | 4 (5.7) | -2.22<br>(-28.57 to 24.13) | 0.866 |
| **Smoker or previous smoker** | 20 (28.6) | 4.40<br>(-6.15 to 14.96) | 0.406 |
| **Obesity** | 23 (32.9) | 3.53<br>(-7.95 to 15.01) | 0.539 |
| **Severity of COVID-19** |  |  |  |
| Mild disease | 42 (60) | ref |  |
| Severe disease | 28 (40) | -13.99<br>(-25.05 to -2.93) | **0.014** |
| **Age groups (years)** |  |  |  |
| 20-59 | 12 (17.1) | ref |  |
| 60-89 | 31 (44.3) | 4.14<br>(-7.58 to 15.87) | 0.481 |
| Multiple linear regression calculated for the dependent variable DL<sub>co</sub> % of predicted (DL<sub>co</sub>%pred). Reported with β-coefficient and 95% confidence interval (CI). DL<sub>co</sub>, diffusion capacity; HA, hyaluronan acid; ref, reference group. Obesity defined as BMI ≥ 30. Adjusted R<sup>2</sup> = 0.304, p < 0.001. Significance set at p < 0.05. | | | |

